# Baseline risk factors associated with immune related adverse events and atezolizumab

**DOI:** 10.1101/2022.11.16.22282410

**Authors:** Katrin Madjar, Rajat Mohindra, Gonzalo Durán-Pacheco, Rashad Rasul, Laurent Essioux, Vidya Maiya, G Scott Chandler

## Abstract

**Background:** Immune checkpoint inhibitors (ICIs) have revolutionized the treatment of cancer patients in the last decade, but immune-related adverse events (irAEs) pose significant clinical challenges. Despite advances in the management of these unique toxicities, there remains an unmet need to further characterize the patient-level drivers of irAEs in order to optimize the benefit/risk balance in patients receiving cancer immunotherapy.

**Methods:** An individual-patient data post-hoc meta-analysis was performed using data from 10,344 patients across 15 Roche sponsored clinical trials with atezolizumab in five different solid tumor types to assess the association between baseline risk factors and the time to onset of irAE. In this study, the overall analysis was conducted by treatment arm, indication, toxicity grade and irAE type, and the study design considered confounder adjustment to assess potential differences in risk factor profiles.

**Results:** This analysis demonstrates that the safety profile of atezolizumab is generally consistent across indications in the 15 studies evaluated. In addition, our findings corroborate with prior reviews which suggest that reported rates of irAEs with PD-(L)1 inhibitors are nominally lower than CTLA-4 inhibitors. In our analysis, there were no remarkable differences in the distribution of toxicity grades between indications, but some indication-specific differences regarding the type of irAE were seen across treatment arms, where pneumonitis mainly occurred in lung cancer, and hypothyroidism and rash had a higher prevalence in advanced renal cell carcinoma compared to all other indications. Results showed consistency of risk factors across indications and by toxicity grade. The strongest and most consistent risk factors were mostly organ-specific such as elevated liver enzymes for hepatitis and thyroid stimulating hormone (TSH) for thyroid toxicities.The main exception to this consistency of risk factors was for ethnicity, which was associated with rash, hepatitis and pneumonitis. Further understanding the impact of ethnicity on ICI associated irAEs is considered as an area for future research.

**Conclusions:** Overall, this analysis demonstrated that atezolizumab safety profile is consistent across indications, is clinically distinguishable from comparator regimens without checkpoint inhibition, and in line with literature, seems to suggest a nominally lower reported rates of irAEs vs CTLA-4 inhibitors. This analysis demonstrates several risk factors for irAEs by indication, severity and location of irAE, and by patient ethnicity. Additionally, several potential irAE risk factors that have been published to date, such as demographic factors, liver enzymes, TSH and blood cell counts, are assessed in this large-scale meta-analysis, providing a more consistent picture of their relevance. However, given the small effects size, changes to clinical management of irAEs associated with the use of Anti-PDL1 therapy are not warranted.

## Introduction

Immune checkpoint inhibitors (ICIs) have revolutionized the care of cancer patients, and these therapies are now the standard of care for treatment in multiple cancer indications. Treatment with ICIs may result in a unique form of toxicity related to the immunological mechanism of action and are commonly referred to as immune-related adverse events (irAEs).

The safety profile of anti-PD(L)1 therapies has been well documented (Brahmer, 2016; Michot, 2016; Postow, 2018; Dougan, 2020;). A recent systematic review found that patients treated with PD-(L)1 inhibitors developed irAEs at a rate of 74% (14% grade ≥3), those treated with CTLA-4 inhibitors at a rate of 89% (34% grade ≥3), and those treated with combination ICIs at a rate of 90% (55% grade ≥3) (Brahmer et al., 2021). Although treatment with ICIs is generally well-tolerated, some events are associated with significant morbidity and mortality (Brahmer, 2018; Postow, 2018; Dougan, 2020; and Morad, 2021). Current guidelines for the clinical management of irAEs clearly demonstrate this severity and call for the development of predictive biomarkers to identify patients at increased risk for such toxicities (Brahmer, 2018; Hussaini, 2020; Brahmer, 2021). Given the morbidity, mortality, and clinical management challenges associated with irAEs, understanding the mechanisms of these immune toxicities and identifying risk factors to predict their development may help to optimize the benefit/risk balance of these therapies and improve patient outcomes. The identification of reliable predictors for risk of irAEs remains an unmet need.

The aim of the current post-hoc meta-analysis was to characterize patient-level drivers of irAEs and seek whether there are any specific baseline factors and/or subpopulations which might be at higher risk of developing ICI associated immune toxicities. Although irAEs may occur in most organ systems, we focused our analysis here on some of the most frequently occurring irAEs (rash, hepatitis, pneumonitis, hypothyroidism and hyperthyroidism) that were reported in 15 Roche sponsored clinical trials with atezolizumab across five different solid tumor types. In these trials, atezolizumab was compared to standard of care or best supportive care, in monotherapy or as part of combination. Using data from these trials, a meta-analysis was conducted to better characterize the profile of irAEs and decipher whether baseline risk factors could help determine potential drivers of irAEs. This study assessed baseline demographic, clinical, laboratory, and tumor characteristics for potential association with irAEs including any organ specificity, or any correlation with severity or differences by tumor type.

This comprehensive meta-analysis based on data from 10,344 patients enrolled in Roche sponsored trials with atezolizumab across five solid tumor indications is one of the largest such analyses to date which provides insight into risk factors for irAEs, an area of ongoing investigation and unmet need.

## Methods

### Patient cohorts

Retrospective meta-analysis of immune-related adverse events (irAE) using individual patient data (all patients in the safety evaluable population) from 15 previously completed randomized controlled phase II or III trials, all sponsored by F. Hoffmann–La Roche/Genentech. These trials tested atezolizumab, either as monotherapy or in combination with chemotherapies and bevacizumab, across five different cancer indications: non-small cell lung cancer (NSCLC) (n=6428; NCT01846416, NCT01903993, NCT02031458, NCT02008227, NCT02409342, NCT02367781, NCT02367794, NCT02657434, NCT02366143), small cell lung cancer (SCLC) (n=494; NCT02763579), urothelial bladder cancer (UC) (n=1331; NCT02951767 & NCT02108652, NCT02302807), advanced renal cell carcinoma (RCC) (n=1201; NCT01984242, NCT02420821), and triple-negative breast cancer (TNBC) (n=890; NCT02425891). Table 1 provides an overview of these trials.

**Table 1:**
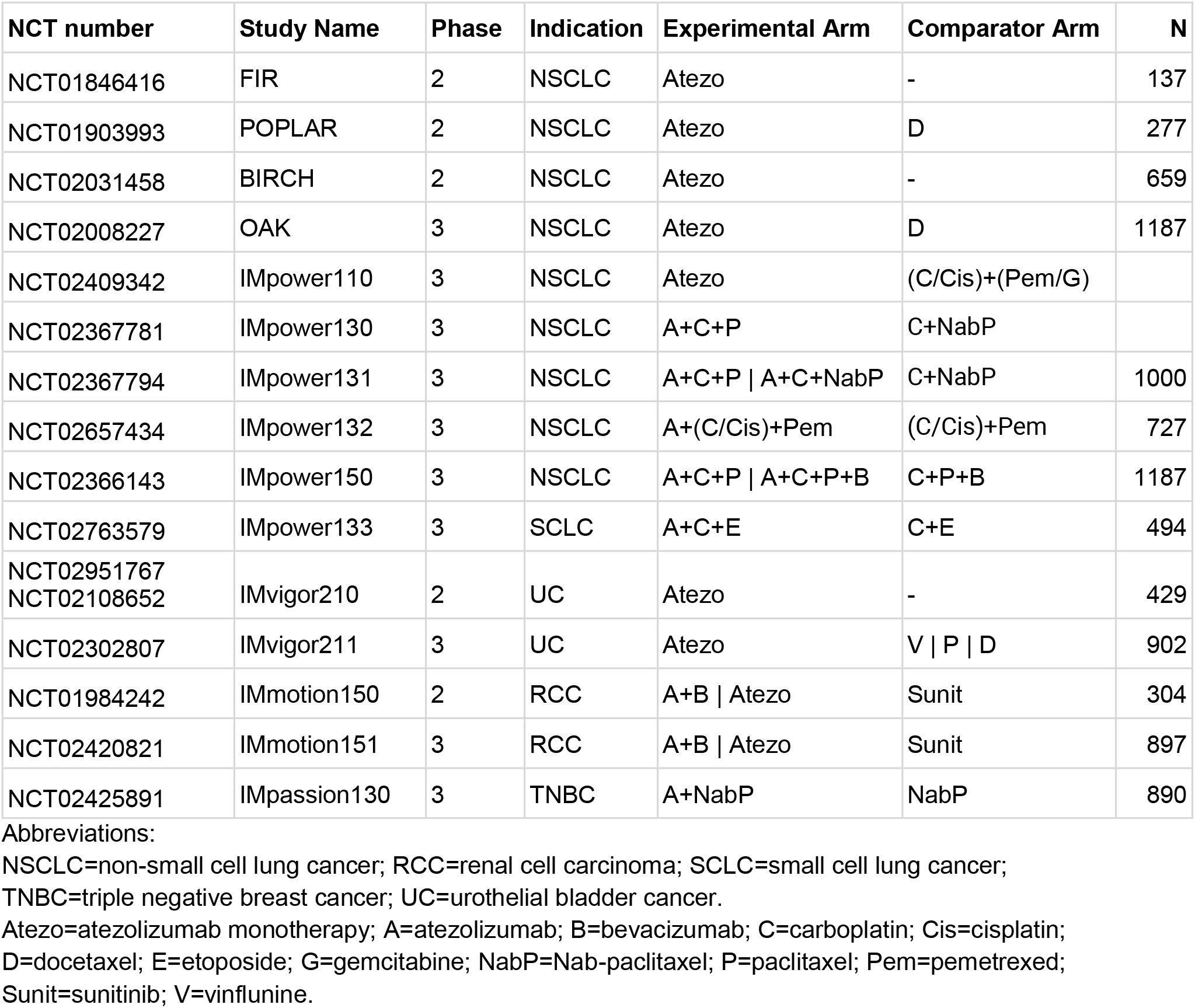
Internal atezolizumab trials with number of patients (N) in the safety evaluable population.

| NCT number | Study Name | Phase | Indication | Experimental Arm | Comparator Arm | N |
| --- | --- | --- | --- | --- | --- | --- |
| NCT01846416 | FIR | 2 | NSCLC | Atezo | - | 137 |
| NCT01903993 | POPLAR | 2 | NSCLC | Atezo | D | 277 |
| NCT02031458 | BIRCH | 2 | NSCLC | Atezo | - | 659 |
| NCT02008227 | OAK | 3 | NSCLC | Atezo | D | 1187 |
| NCT02409342 | IMpower110 | 3 | NSCLC | Atezo | (C/Cis)+(Pem/G) |  |
| NCT02367781 | IMpower130 | 3 | NSCLC | A+C+P | C+NabP |  |
| NCT02367794 | IMpower131 | 3 | NSCLC | A+C+P \| A+C+NabP | C+NabP | 1000 |
| NCT02657434 | IMpower132 | 3 | NSCLC | A+(C/Cis)+Pem | (C/Cis)+Pem | 727 |
| NCT02366143 | IMpower150 | 3 | NSCLC | A+C+P \| A+C+P+B | C+P+B | 1187 |
| NCT02763579 | IMpower133 | 3 | SCLC | A+C+E | C+E | 494 |
| NCT02951767<br>NCT02108652 | IMvigor210 | 2 | UC | Atezo | - | 429 |
| NCT02302807 | IMvigor211 | 3 | UC | Atezo | V \| P \| D | 902 |
| NCT01984242 | IMmotion150 | 2 | RCC | A+B \| Atezo | Sunit | 304 |
| NCT02420821 | IMmotion151 | 3 | RCC | A+B \| Atezo | Sunit | 897 |
| NCT02425891 | IMpassion130 | 3 | TNBC | A+NabP | NabP | 890 |

Abbreviations:
NSCLC=non-small cell lung cancer; RCC=renal cell carcinoma; SCLC=small cell lung cancer;
TNBC=triple negative breast cancer; UC=urothelial bladder cancer.
Atezo=atezolizumab monotherapy; A=atezolizumab; B=bevacizumab; C=carboplatin; Cis=cisplatin;
D=docetaxel; E=etoposide; G=gemcitabine; NabP=Nab-paclitaxel; P=paclitaxel; Pem=pemetrexed;
Sunit=sunitinib; V=vinflunine.

### Endpoints

For each patient, the onset of the first irAE of a specific type (rash, hepatitis, pneumonitis, hypothyroidism or hyperthyroidism) was considered, i.e. for a specific irAE type every patient was counted only once even if the patient had more than one event of the same type. The irAEs selected for this analysis are usually reported most commonly with ICIs i.e. rash, hepatitis (diagnosis and laboratory abnormalities), pneumonitis, hypothyroidism and hyperthyroidism.

The safety time-to-event endpoint was defined by the time from the first treatment administration to the onset of the first irAE or censoring due to death, completion or discontinuation, or end of AE reporting period. In the latter case, the censoring time was defined as the last treatment date plus 30 days.

The CTCAE toxicity grade was accounted for by separately analyzing the first irAE of any grade, of grade 2 or higher (2+), and of grade 3 or higher (3+). If not stated differently, the first irAE is referenced irrespective of the toxicity grade (any grade).

The present analyses did not account for different lengths of safety follow-up times between arms (e.g. in studies POPLAR, IMpower131, IMpassion130 and IMvigor211). However, to assess the impact on the risk factor estimates another analysis was performed, using the study-specific publication data cuts to align the follow-up periods between arms. The results were compared to the results presented in this publication, showing consistency, i.e. that the risk factor estimates were not affected by the different follow-up lengths (data not shown).

### Adverse events

Adverse events were captured per the criteria specified in the study protocols. For the purpose of analysis of immune-related AEs, a set of comprehensive definitions comprising Sponsor defined, Standardized Medical Dictionary for Regulatory Activities (MedDRA) Queries (SMQ), High Level Terms (HLTs), and Sponsor defined AE Grouped Terms (AEGTs) were used to identify and summarize irAEs by medical concept. The definitions used to capture the medical concepts were broad in nature and were applied to capture AEs both in atezolizumab and standard of care arms irrespective of causality. The medical concepts represented are identified risks, potential risks or class effects associated with immune checkpoint inhibitors. NCI CTCAE criteria were used for the severity grading of AEs and immune-related AEs.

### Data preprocessing

Numeric variables (tested risk factors and the two confounders of age and body mass index [BMI]) were preprocessed before performing the meta-analyses by identification and exclusion of extreme values (values that were 15 fold greater/smaller than the upper/lower boxplot whisker defined by the quantiles method implemented in the R ‘boxplot’ function), natural log-transformation of values (values ≤0 were set to 0.01 before log-transformation) and standardization to mean 0 and standard deviation (SD) of 1. Consequently, the resulting hazard ratios (HR) had the same scale enabling a direct comparison of effect sizes between different covariates. The HR could be interpreted as the change in the hazard of irAE onset per change in 1 SD unit at the log scale of a given covariate.

Missing values were not imputed. Missing data were discarded from the analysis and a complete case analysis was performed.

For laboratory parameters all available baseline data were considered in the analysis (e.g. not only elevated levels).

### Competing event

When a patient died within the safety follow-up period it was considered a competing event since its occurrence stopped the safety follow-up for that patient and prevented any potential future irAE from happening.

To obtain an unbiased estimate of the irAE probability over time, death was accounted for as a competing event using the non-parametric Aalen-Johansen (AJ) estimator of the cumulative incidence function (CIF). CIF plots based on the AJ estimator and irAE frequencies per study/indication and treatment arm are used for a descriptive comparison of the toxicity profiles. The advantage of the AJ estimator over simple frequencies is that it accounts for censoring, different lengths of follow-up periods (e.g. across treatment arms), and competing events.

In the two meta-analysis approaches death was considered as a competing event for the irAE of interest by using a Cox proportional cause-specific hazards regression model. This cause-specific approach treats competing events (here: death) as censored observations (Stegherr et al., 2021; Wolbers et al., 2014).

### Identification of baseline risk factors

Baseline risk factors (demographics, clinical, laboratory parameters, and tumor characteristics) were tested for an association with the onset of irAEs based on the aggregated patient-level data from 15 trials using an individual-patient data (IPD) meta-analysis. Each irAE medical concept (rash, hepatitis, pneumonitis, hypothyroidism and hyperthyroidism) was considered separately. The data was adjusted for the following important confounders in the model: age (years), sex (female vs male), BMI (kg/m^2^) and treatment (atezo-mono, atezo-combo, standard of care), and multiple testing correction was applied to the list of tested factors.

The meta-analysis was conducted across all indications and separately in each indication (lung cancer [combining NSCLC and SCLC], UC and RCC), as well as across different treatment arms and separately in the atezolizumab arms (monotherapy or combination) and in the standard of care arms (SOC, chemotherapy or targeted therapy). Sub-analysis included analysis by irAE severity and comparing the results for irAEs of any grade with those of higher grade (2+ or 3+).

### Individual patient data meta-analysis

To assess the association of baseline risk factors with the time to onset of irAE, a mixed-effects Cox proportional hazards regression model (implemented in the R package ‘coxme’ version 2.2-16) was used and adjusted for the selected confounders age (years), sex (female vs male), BMI (kg/m^2^) and treatment (atezo-mono, atezo-combo, SOC). The model equation is

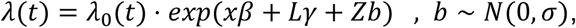

where λ_0_(*t*) is the baseline hazard, *x* is the baseline risk factor of interest and *β* the corresponding fixed-effects coefficient, *L* is the design matrix for the fixed-effects of the confounding covariates and *γ* is the corresponding fixed-effects coefficient vector, *Z* is the design matrix for the study random-effects and *b* is the corresponding random-effects coefficient vector.

The risk factors of interest *x* were tested independently by adding one at a time to the model. Their prognostic effect was assessed by hazard ratios (HR) *exp*(*β*) and corresponding 95%-confidence intervals (CI), as well as Wald test p-values for hypothesis testing of *β*. The p-values were adjusted for multiple testing using the Benjamini & Hochberg’s (1995) FDR correction and the analysis was conducted using separate multiple testing correction for each irAE medical concept and each analysis.

### Study-level meta-analysis

In a first step, a separate Cox proportional hazards regression model was fitted to each study population including the baseline risk factor of interest *x* and adjusting for important confounders (age, sex, BMI, treatment).

In a second step, the study-level effects *β*_*i*_of the risk factor *x* (log HR and corresponding standard errors for each study) were combined using a random-effects meta-analysis to estimate the pooled effect *β* across all studies as follows:

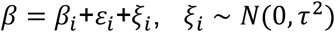

where *β*_*i*_ is the log HR of the risk factor for study *i, ε*_*i*_and *ξ*_*i*_are the within and between study residuals respectively. *τ*^2^ is the variance of the distribution of study-level effects or heterogeneity parameter. The Sidik-Jonkman’s method was used to estimate the between-study variance (*τ*^2^) and the Hartung-Knapp-Sidik-Jonkman method to estimate the variance of the meta-analysis overall effect *var*(β). The random-effects meta-analysis was performed with the R package ‘meta’ (version 4.11-0).

## Results

The association between baseline risk factors (demographics, clinical, laboratory parameters, and tumor characteristics) and the onset of specific irAEs was examined based on patient-level data from 10,344 patients from 15 internal clinical phase II or III trials testing atezolizumab, either as monotherapy or in combination with chemotherapies and bevacizumab, across five different cancer indications (NSCLC, SCLC, UC, RCC, TNBC) (Table 1).

An overview of patient characteristics across and by indication and treatment arm is provided to understand whether there are differences in the safety risk factor profile by indication or between atezolizumab and standard of care. The timing, type (location) and severity of irAEs was also considered.

### Toxicity profile by indication and treatment

Table 2 shows aggregated irAEs for the five irAE types of interest across all trials stratified by the indication and treatment arm. For each patient, only the first irAE of a specific type irrespective of the severity was considered. The most prevalent irAEs (all grades) were rash (22.77%) and hepatitis (12.35%), while hypothyroidism (8.96%), pneumonitis (3.01%), and hyperthyroidism (2.42%) occurred less frequently, which is in accordance with the most commonly affected organ systems reported in the literature (Manson et al., 2016; Postow et al., 2018). The frequency of irAEs was higher under atezolizumab (either monotherapy or combination) than under chemotherapy, as expected based on mechanism of actions of immune checkpoint inhibitors, and lower as compared to sunitinib (targeted therapy in RCC) for rash, hepatitis and hypothyroidism.

**Table 2:** Frequencies of irAEs (rash, hepatitis, pneumonitis, hypothyroidism, hyperthyroidism) of any grade per indication and treatment arm (atezolizumab monotherapy, atezolizumab combination, standard of care/control arms [chemotherapy or targeted therapy]). For each patient only the first irAE of each type was considered.

| Indic. | Treatment | Rash | Hepatitis | Pneumonitis | Hypothyroid. | Hyperthyroid. | N |
| --- | --- | --- | --- | --- | --- | --- | --- |
| Lung C. | Atezo-mono | 350 (19.12%) | 206 (11.25%) | 68 (3.71%) | 112 (6.12%) | 29 (1.58%) | 1831 |
|  | Atezo-combo | 669 (26.21%) | 418 (16.38%) | 155 (6.07%) | 309 (12.11%) | 104 (4.08%) | 2552 |
|  | Chemo | 371 (14.61%) | 216 (8.51%) | 34 (1.34%) | 51 (2.01%) | 25 (0.98%) | 2539 |
| RCC | Atezo-mono | 40 (38.83%) | 10 (9.71%) | 1 (0.97%) | 24 (23.3%) | 2 (1.94%) | 103 |
|  | Atezo-combo | 193 (34.96%) | 74 (13.41%) | 13 (2.36%) | 141 (25.54%) | 34 (6.16%) | 552 |
|  | Sunit | 317 (58.06%) | 115 (21.06%) | 0 (0%) | 169 (30.95%) | 22 (4.03%) | 546 |
| UC | Atezo-mono | 181 (20.38%) | 94 (10.59%) | 20 (2.25%) | 42 (4.73%) | 14 (1.58%) | 888 |
|  | Chemo | 49 (11.06%) | 26 (5.87%) | 3 (0.68%) | 0 (0%) | 0 (0%) | 443 |
| TNBC | Atezo-combo | 98 (22.02%) | 62 (13.93%) | 15 (3.37%) | 64 (14.38%) | 15 (3.37%) | 445 |
|  | Chemo | 87 (19.55%) | 57 (12.81%) | 2 (0.45%) | 15 (3.37%) | 5 (1.12%) | 445 |
| **Total** |  | **2355 (22.77%)** | **1278 (12.35%)** | **311 (3.01%)** | **927 (8.96%)** | **250 (2.42%)** | **10344** |

We further examined the severity of each type of irAE and found that the majority of irAEs were of grade 1, except for hypothyroidism and pneumonitis where grade 2 was most frequently reported. We observed grade 3 and 4 irAEs mainly for hepatitis and pneumonitis, the frequency of grade 5 irAEs was very low. Overall the distribution of toxicity grades is consistent across indications, however, there are small indication-specific differences in the irAE type which are irrespective of the treatment: pneumonitis mainly occurred in lung cancer, whereas hypothyroidism and rash had a higher prevalence in RCC compared to all other indications (Figure 1).

**Figure 1:**
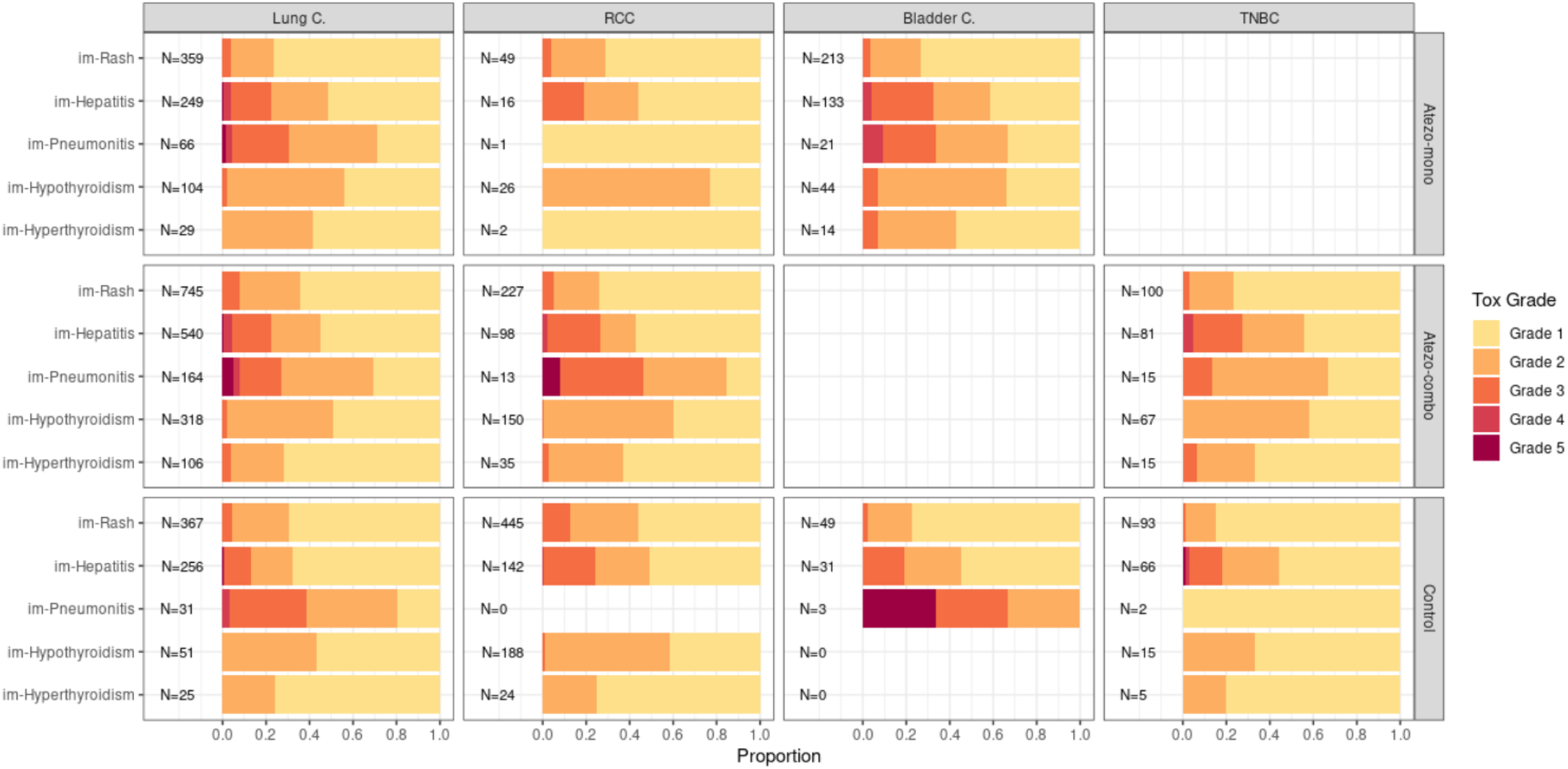
For the patients with irAEs the proportion of irAEs by toxicity grade are shown for each treatment arm (row panels) and indication (column panels), together with the total number of events per arm and indication (N). Toxicity grade data is not available for the POPLAR study (NSCLC).

Supplementary Figure 1 shows the cumulative incidence for each type of irAE over time, taking into account censoring, varying follow-up periods across studies and arms, timing of the events, and additionally considering death as a competing event in order to ensure an unbiased estimation of the irAE probability. The irAE probabilities under atezolizumab (either monotherapy or combination) tended to be higher compared to chemotherapy and lower compared to sunitinib in RCC. There was no clear difference in irAE risk with and without bevacizumab added to either atezolizumab alone (IMmotion150 study) or atezolizumab plus chemotherapy (IMpower150 study). The remaining studies had either atezolizumab monotherapy or a combination of atezolizumab and therefore, did not allow a direct comparison of the two arms.

### Identification of baseline risk factors

The confounder-adjusted IPD meta-analysis results for each irAE type and all different analyses are summarized in Supplementary Table 1. These findings are visualized in Figure 2 in terms of effect size estimates (hazard ratios) and significance levels for all tested baseline risk factors with an FDR-adjusted p-value <0.1 in at least one of the analyses.

**Figure 2:**
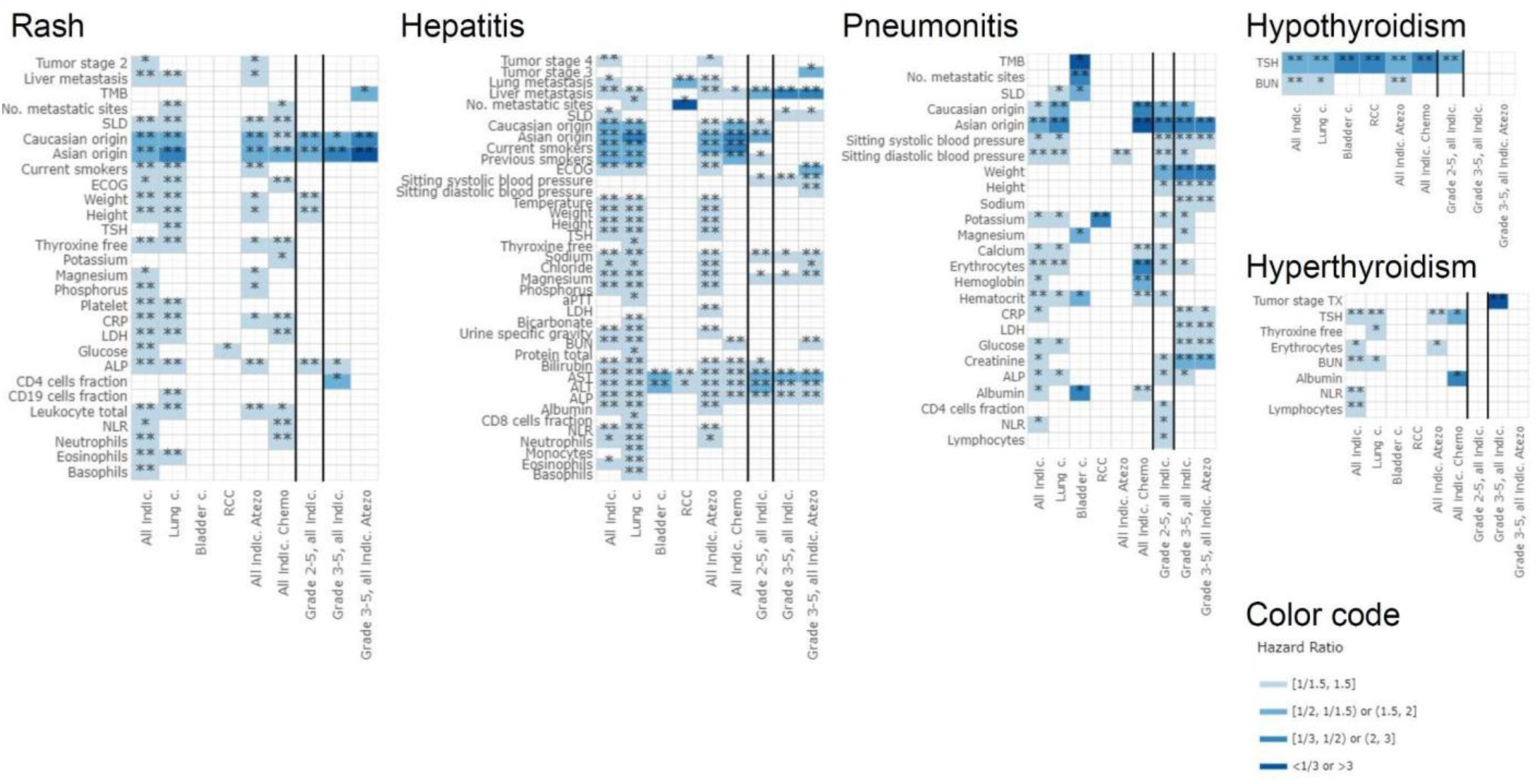
Effect estimates from the IPD meta-analysis for the baseline risk factors (rows) with an FDR-adjusted p-value <0.1 in at least one of the analyses (columns). The color refers to the absolute hazard ratio size representing the strength of association. The symbol refers to the level of significance (FDR p-value <0.1: *, FDR p-value <0.05: **). The following different analyses were performed: “All indic.” (across different indications and arms), “Lung c.” (only lung cancer studies including all arms), “Bladder c.” (only UC studies including all arms), “RCC” (only RCC studies including all arms), “All indic. Atezo” (across different indications but only atezolizumab arms), “All indic. Chemo” (across different indications but only standard of care arms), “Grade 2-5, all indic.” (across different indications and arms, only grade 2+ irAEs), “Grade 3-5, all indic.” (across different indications and arms, only grade 3+ irAEs), “Grade 3-5, all indic. Atezo” (across different indications, only atezolizumab arms and grade 3+ irAEs). Abbreviations of risk factors: ALP=Alkaline Phosphatase, ALT=Alanine Aminotransferase, aPTT=activated Partial Thromboplastin Time, AST=Aspartate Aminotransferase, BUN=Blood Urea Nitrogen, CRP=C-Reactive Protein, ECOG=Eastern Cooperative Oncology Group performance status, LDH=Lactate Dehydrogenase, NLR=Neutrophil-Lymphocyte Ratio, SLD=Sum of the Longest Diameters, TMB=Tumor Mutational Burden, TSH=Thyroid Stimulating Hormone.

One of the strongest potential risk factors that consistently came up was the association of Asian ancestry (based on self-reported ethnicity) with the development of rash (hazard ratios ranging from 1.74 to 2.03 for any [mainly lower] grade rash and from 2.00 to 3.39 for higher grade [2+ or 3+]); hepatitis (hazard ratios ranging from 1.58 to 2.21); and pneumonitis (hazard ratios ranging from 1.87 to 3.97). The direction of the effect was consistent with patients reporting Asian origin exhibiting a higher risk for the development of irAEs (rash, hepatitis, or pneumonitis) than non-Asians, except for RCC (patients of Asian origin having a lower risk of hepatitis and pneumonitis, but this effect was not significant).

Given that Asian origin was one of the strongest risk factors that was identified under all treatment arms, we sought to understand whether the elevated irAE risk in Asian patients was due to country-specific differences in reporting or potentially caused by underlying ethnic differences such as genetic factors. In the combined cohort all patients reported in Asian countries (n=397 from China and Taiwan, n=680 from Japan, n=289 from Korea, n=128 from Southeast Asia) were of Asian ancestry except for two Caucasians from Southeast Asia (based on self-reported race). Therefore, we compared the irAE risk for Asians from the different Asian countries with the irAE risk for Asians from all non-Asian countries (non-Asian countries were grouped together for sample size reasons). Figure 3 summarizes the corresponding results of this country of origin effect in Asians. Patients from all Asian countries showed a (numerically) increased risk of hepatitis (China and Taiwan: HR 4.28, 95%-CI [2.34; 7.83]; Japan: HR 2.98, 95%-CI [1.64; 5.44]; Korea: HR 1.38, 95%-CI [0.68; 2.81]; Southeast Asia: HR 2.10, 95%-CI [1.01; 4.38]) and pneumonitis (China and Taiwan: HR 1.91, 95%-CI [0.54; 6.81]; Japan: HR 3.12, 95%-CI [0.95; 10.23]; Korea: HR 2.10, 95%-CI [0.53; 8.25]; Southeast Asia: HR 2.68, 95%-CI [0.66; 10.86]) compared to the patients from non-Asian countries. However, for rash the risk was higher only in Japan (HR 1.46, 95%-CI [1.06; 2.02]) and lower in all other Asian countries (China and Taiwan: HR 0.65, 95%-CI [0.45; 0.96], Korea: HR 0.74, 95%-CI [0.49; 1.10], Southeast Asia: HR 0.86, 95%-CI [0.54; 1.38]). This effect was independent on whether confounder adjustment was performed on age, BMI, sex and treatment or not, thus, indicating a potential reporting bias for skin toxicities in Japan.

**Figure 3:**
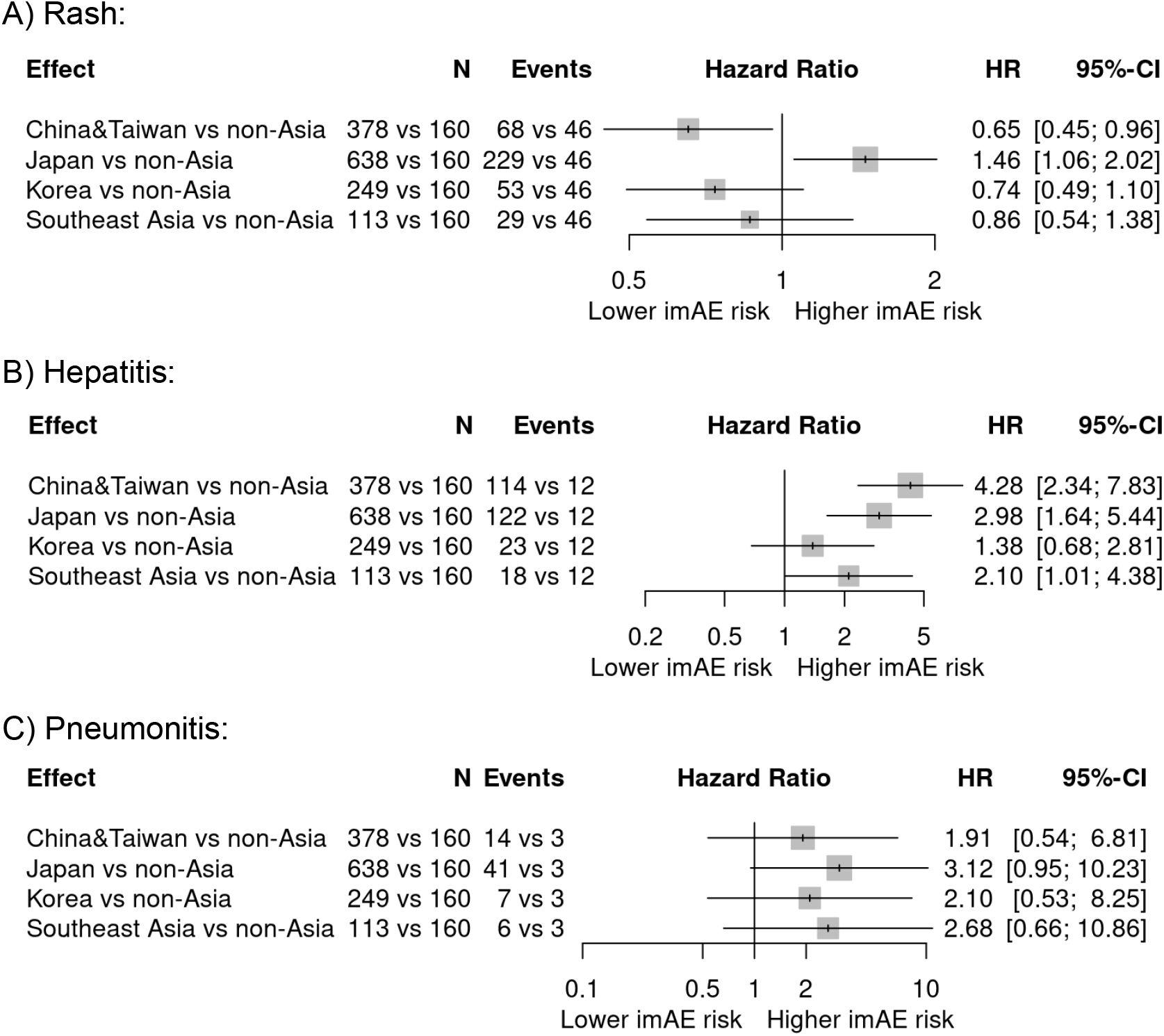
Forest Plots of the country effect in patients with Asian ancestry (hazard ratio [HR] estimates and 95%-confidence intervals [CI]) adjusted for important confounders: age, BMI, sex and treatment. Each Asian country is compared against the group of all non-Asian countries (reference group). The effect estimates are obtained by combining data from all Asian patients across all studies in an individual patient data meta-analysis including country and the confounding factors as fixed effects and indication as a random effect. The sample size (N) and number of irAEs (Events) are given for patients belonging to the specific Asian country vs those who are from the reference group of non-Asian countries (these are the actual numbers used for the analysis, i.e. excluding patients with missing values in any of the above mentioned variables included in the model).

Other important demographic factors, such as age, sex, and BMI, were included as confounders in every risk factor model. Higher BMI values were consistently associated with an increased risk of rash (HR 1.09, 95%-CI [1.04; 1.13]), younger patients were more likely to develop hepatitis (HR 0.81, 95%-CI [0.77; 0.85]), and females had a higher risk of hypothyroidism (HR 1.37, 95%-CI [1.18; 1.60]) and a lower risk of pneumonitis (HR 0.66, 95%-CI [0.50; 0.86]) as compared to males (Supplementary Figure 2 and Supplementary Table 2).

Further important risk factors in the overall population were baseline elevations in liver enzymes (ALT, AST, ALP) (hazard ratios ranging from 1.26 to 1.69) and liver metastasis (hazard ratios ranging from 1.38 to 2.07) for the association with hepatitis, where higher baseline values of liver enzymes and presence of liver metastasis were associated with an increased hepatitis risk. Elevated baseline TSH levels were associated with a higher risk of hypothyroidism (hazard ratios ranging from 1.62 to 2.57) and a lower risk of hyperthyroidism (hazard ratios ranging from 0.66 to 0.77).

Blood cells, such as Neutrophil-Lymphocyte Ratio (NLR) or Platelet counts, have been reported in the literature as risk factors for irAEs. We found that Platelet count was associated only with a slight decreased risk of rash (HR=0.94), whereas NLR came up as a protective factor with decreased risk of rash (HR=0.93), hepatitis (HR=0.89) and hyperthyroidism (HR=0.77), and as a risk factor with increased risk of pneumonitis (HR=1.19). In an additional analysis we assessed whether the combination of NLR and Platelet, either included as two separate risk factors in the model or combined in the new risk factor systemic immune-inflammation index (SII) (NLR x Platelet), would strengthen the association with the five irAEs of interest. SII was only associated with a slight decreased hepatitis risk (HR=0.93). When combining the two separate risk factors NLR and Platelet, only the effect of NLR remained similar (significance level and effect estimate). Thus, NLR is the dominant risk factor and the combination of NLR and Platelet does not add any prognostic value to NLR (Supplementary Table 3).

C-Reactive Protein (CRP) was associated with a decreased risk of rash of any grade (mainly G1), and an increased risk of grade 3 or higher pneumonitis irrespective of the treatment (under both atezolizumab and SOC) (rash: HR=0.81-0.92, pneumonitis: HR=1.47-1.48). We found a very similar relationship with the irAEs of interest for LDH (rash of any grade: HR=0.91-0.93, pneumonitis of grade 3+: HR=1.38-1.42), and SLD (only rash of any grade: HR=0.86-0.91). For all three risk factors (CRP, LDH, SLD) the association with rash was slightly more pronounced under SOC as compared to atezolizumab.

Tumor mutational burden (TMB) was not associated with the selected irAEs of interest, which is in line with published results (Wells et al., 2017; Osipov et al., 2020).

A similar analysis was performed using a study-level (SL) meta-analysis approach and the results of the risk factors (hazard ratios and confidence intervals) were compared to those from the IPD meta-analysis (results not shown). The results were consistent across the two different statistical approaches with hazard ratios being highly correlated. Only the confidence intervals from the SL meta-analysis approach tended to be larger due to the smaller sample size.

Overall the findings were relatively consistent across indications (lung cancer, UC and RCC) and treatment arms (atezolizumab vs SOC).The sample size in UC and RCC was relatively small, making interpretation of results difficult, however the directionality of the findings was similar to the overall analysis. In general, the identified risk factors were not unique to atezolizumab but were also associated with AEs in the SOC arms. The broad definitions used to capture the irAE medical concepts likely explains the consistency of the findings across the treatment arms. The similar associations also point to the absence of treatment-specific predictive factors among those factors investigated (Supplementary Figure 3).

With regard to irAE severity, we also obtained consistent results for any grade as compared to higher grade (2+ or 3+) with a few additional risk factors related to higher grade pneumonitis.

There were no common risk factors shared by all five irAE types, suggesting that risk factors were rather organ-specific except for the self-reported ethnic background.

## Discussion

Treatment with ICIs has revolutionized cancer therapy in the last decade since their first approval. However, the irAEs associated with these treatments pose unique clinical management challenges compared to other cancer therapies (Brahmer, 2018; Postow, 2018; Dougan, 2020; and Brahmer, 2021). Although these irAEs are currently understood to be the sequelae of the intended restoration of immune function following ICI therapy, the underlying host-specific and tumor-specific factors have yet to be fully determined (Chen, 2017; Postow, 2018; Morad, 2021). Despite efforts to further characterize the underlying mechanisms driving these toxicities, reliable biomarkers and predictive risk factors have yet to be identified (Brahmer, 2018; Brahmer, 2021).

In this large post-hoc meta-analysis, including 10,344 patients from 15 atezolizumab clinical trials across five indications, we provide a comprehensive view of baseline risk factors associated with occurrence of irAE in patients treated with atezolizumab monotherapy and combinations, and in comparison to standard of care chemotherapy or targeted therapy. To our knowledge, such a large-scale analysis of safety risk factors has not been conducted to date. Reported baseline risk factors in the literature are usually based on analysis of a collection of various types of irAEs, either for one specific indication or combining different indications, and focussing on ICI treatment without comparing to standard of care.

In general, the safety profile of atezolizumab was consistent across the five indications presented here (NSCLC, SCLC, UC, RCC, and TNBC) except for some small differences, i.e. higher frequency of pneumonitis in lung cancer and higher frequency of hypothyroidism and rash in RCC (Table 2). Overall, 44.77% of the patients treated with atezolizumab experienced at least one irAE (all grade) and 9.29% had grade 3 or higher. The frequency of irAEs was higher under atezolizumab (either monotherapy or combination) than under chemotherapy, as expected based on mechanism of actions of immune checkpoint inhibitors, and lower as compared to sunitinib (targeted therapy in RCC) for rash, hepatitis and hypothyroidism. Overall, our analysis suggests that the reported rates of irAEs with PD-(L)1 inhibitors is nominally lower than CTLA-4 inhibitors as cited in literature. A recent review comparing the safety profiles across multiple ICI regimens demonstrated that patients treated with PD-(L)1 inhibitors developed irAEs at a rate of 74% (14% grade ≥3), those treated with CTLA-4 inhibitors at a rate of 89% (34% grade ≥3), and those treated with combination ICIs at a rate of 90% (55% grade ≥3 [Brahmer, 2021]). This recent review was in concordance with publication from Michot et al. (2016) that highlighted the frequency of irAEs as upto 90% with CTLA-4 inhibitors and 70% in patients treated with PD-1/PD-L1 antibodies.

The most prevalent irAEs in this analysis (all grades) were rash (22.77%), hepatitis (12.35%), and hypothyroidism (8.96%), while pneumonitis (3.01%), and hyperthyroidism (2.42%) were relatively less common. This is in accordance with the most commonly affected organ systems reported in the literature (Manson et al., 2016; Postow et al., 2018) and prevalence of irAEs with atezolizumab appear nominally lower than CTLA-4 inhibitors (Michot et al., 2016).

On average, the time to onset of a patient’s first irAE for the five selected irAEs of interest was 10.4 weeks (median). The irAE with shortest median time to onset was rash with 6.3 weeks. The median time to resolution for these selected irAEs of interest was 3.6 weeks (i.e. irAE duration using the patients’ first irAEs).

In this analysis, several candidate baseline risk factors, such as self-reported ethnicity, TSH, and liver enzymes, were associated with the onset of irAE in the treatment arms. The apparent similarity in associations between treatment arms may point to the absence of treatment-specific predictive factors among those factors investigated. However, the broad definitions used to capture the irAE medical concepts and the blinded nature of studies at the time when safety events were reported likely confound the irAE baseline risk factor related interpretation in the standard of care arms. Although these findings may hold potential predictive value for identifying those at risk of developing such irAEs, the effect size of each of these associations was relatively modest, and do not warrant changes to clinical management at this time. Furthermore, we did not find differences between lower and higher irAE toxicity grades nor were any indication-specific risk factors identified. However, as would be expected clinically, certain organ specific risk factors were identified, including increased liver enzymes associated with on-treatment hepatitis and abnormal TSH at baseline associated with on-treatment thyroid irAEs. Additionally, there were some differences identified in important demographic factors such as sex and ethnicity (Figure 2).

## Demographic factors

**Ethnicity** is emerging as one of the most important demographic factors with respect to cancer diagnosis, prognosis, and the impact of healthcare disparities. These disparities in cancer incidence and outcomes by race/ethnicity result from the interplay between structural, socioeconomic, socio-environmental, behavioral and biological factors. In our meta-analysis, ethnic origin was the only relatively strong and consistent risk factor that was identified across multiple irAEs, with Asians being at higher risk of rash, pneumonitis and hepatitis.

Thompson et al. (2021) suggested differences in reporting of cutaneous irAEs (cirAE) between white and non-white patients (about 51% of the non-white patients being Asians and 30% being African Americans). The authors observed that non-white patients were half as likely to be diagnosed with a cirAE and nearly six times more likely to be referred to a dermatology specialist for cirAE management than white patients. Similar country-specific-reporting differences might have confounded the interpretation in our meta-analysis as illustrated by the inconsistent rash risk between Asians from different countries of origin. Specifically, the data suggested that patients of self-reported Asian ancestry in Japan were at higher risk than patients of self-reported Asian ancestry outside Asian countries, whereas in other Asian countries the effect was going in the opposite direction, independent of confounder adjustment (Figure 3).

Asian ethnicity has also been recognized as a potential risk factor of ICI associated pneumonitis (Naidoo et al., 2020). Based on data from use of ICIs in patients with lung cancer, the incidence and discontinuation rate due to ICI associated pneumonitis seemed to be higher in Japanese patients vs other Asian countries. Given the high incidence of interstitial lung disease associated with EGFR TKI in the Japanese population compared with Caucasians and other Asian countries, it is possible that there might be differences in patterns of ICI associated pneumonitis between Japan and other Asian countries (Lee et al., 2020).

The incidence of ICI associated hepatotoxicity of up to 18% has been seen in some studies in Chinese patients and is an important consideration given the higher proportion of hepatitis B virus infections in Asian patients (Peng et al., 2018).

Overall, there are outstanding scientific questions to characterize the impact of ethnicity on ICI associated immune toxicities and further research is needed to progress inclusive research in order to optimize health outcomes for all patients worldwide. Pharmaceutical companies have publicly committed to advancing inclusive research and delivery of health outcomes.

**Age** is an important demographic factor in cancer patients. Data from NCI’s Surveillance, Epidemiology, and End-Results (SEER) program demonstrate that between 2014-2018, more than half of cancers were reported in patients 65 years of age or older. Despite the known impact of aging on immune function, and especially susceptibility to cancer, evidence for the safety and efficacy of ICIs in older patients is limited and mostly derived from subgroup analyses and meta-analyses from clinical trials (Bhandari, 2018). Nevertheless, ICIs are generally well-tolerated in older patients and have a similar overall safety profile (Bhandari, 2018; Nishijima, 2016; Friedman, 2017; Helissey, 2016). Similarly, our analysis showed no specific association between age and the rate or severity of irAEs except for rash and hyperthyroidism where the association was not strong. Although the majority of evidence indicates that there is no difference in the rate and severity of irAEs in older patients, the clinical management of older patients experiencing toxicity from cancer therapies poses additional challenges compared to younger patients (Bhandari, 2018; Extermann, 2006).

**Sex** is another important demographic factor with respect to risk of developing cancer, dying from cancer, and ICI tolerability. While females have a slightly lower risk of developing cancer, as well as dying from cancer, compared to males, females have been reported to be at a higher risk of several autoimmune diseases (Klein and Flanagan 2016; Schwinge and Schramm, 2018). However, the association between sex and irAEs remains unclear with conflicting results reported in the literature. Jing et al. (2021) performed a meta-analysis on published clinical study data with anti-PD-1 or anti-PD-L1 agents and demonstrated no statistically significant irAE risk difference between males and females. The findings from our meta-analysis demonstrate a higher risk of hypothyroidism and a lower risk of pneumonitis in female compared to male patients (Supplementary Figure 2 and Supplementary Table 2), but does not necessarily contradict the published literature as those publications considered combined irAEs in general.

**Body Mass Index (BMI)** is emerging as another potential predictive factor for irAEs due to the proinflammatory state associated with obesity and its association with autoimmune disorders (Guzman-Prado, 2021; Harpsoe, 2014; and Versini, 2019). We observed that higher body mass index (BMI) values were associated with an increased risk of rash (Supplementary Figure 2 and Supplementary Table 2). This relationship was reported for PD-1-related irAEs by one study (Eun et al., 2019), but not by other studies in patients undergoing anti-PD-1, anti-CTLA-4 or anti-PD-L1 therapies (Kartolo et al., 2018; Richtig et al., 2018).

### Clinical/laboratory parameters

Monitoring of liver function tests (LFTs) are critical in the diagnosis and clinical management of patients with ICI-induced hepatotoxicity. However, the potential for liver involvement with the tumor (both primary or metastatic, baseline or on-treatment) add complexity in the interpretation of baseline and on-treatment LFTs, and the corresponding impact of the tumor involvement of the liver, ICI-mediated hepatotoxicity, or both. Although most patients with significant liver disorders at baseline were excluded from the clinical trials in this analysis; patients with stable liver metastasis were included (i.e. excluding patients with baseline AST, ALT and ALP >2.5 x upper limit of normal range [ULN], or patients with baseline liver metastasis and AST, ALT, ALP >5 x ULN). This is important given that some cases of severe immune-mediated hepatotoxicity have been reported in patients with clinically silent liver metastases (Jennings, 2019; Suzman, 2018). In our meta-analysis, baseline elevation in liver enzymes and presence of liver metastasis were associated with an increased risk of hepatitis (Figure 2 and Supplementary Table 1).

Some serum chemistry parameters and measurements of specific organ function have been associated with the pathophysiology of irAEs, and are also important in the diagnosis and management of specific irAEs. ICI-associated thyroiditis is a distinct clinical entity and its mechanistic basis is not well understood (Muir et al., 2020). Baseline thyroid function has emerged as a potential biomarker of response to ICI therapy. The typical pattern of immune-mediated thyroid dysfunction associated with ICI therapy is an initial hyperthyroidism followed by eventual hypothyroidism due to permanent destruction of the gland and the need for potentially lifelong thyroid replacement (Muir, 2020; Olsson-Brown, 2020). Multiple studies have demonstrated a correlation between elevated TSH at baseline and increased risk of anti-PD(L)-1-induced thyroid dysfunction (mainly thyrotoxicosis followed by hypothyroidism) in solid tumors (Kimbara et al., 2018; Ma et al., 2019; Osorio, 2017; Muir, 2021). In our analysis, elevated TSH levels at baseline were consistently associated with a lower risk of hyperthyroidism and a higher risk of hypothyroidism (Figure 2 and Supplementary Table 1). Additionally, our findings showed an increased risk of hypothyroidism in females as compared to males (Supplementary Figure 2 and Supplementary Table 2). For hyperthyroidism the direction of this effect was the same but the effect size was smaller.

Several blood cell parameters have been reported to be associated with the development of irAEs. In particular, the neutrophil-to-lymphocyte ratio (NLR) and the systemic immune-inflammation index (SII) have been associated with increased risk of irAE, as well as improved response; however, there are conflicting reports regarding this association. In our meta-analyses, we did not find strong or consistent associations between the different types of blood cell counts, such as basophils, eosinophils, lymphocytes, monocytes, neutrophils, NLR, leukocytes, erythrocytes and platelets, measured at baseline and the onset of irAEs (see Figure 2 and Supplementary Table 1). NLR was associated with a decreased risk of rash, hepatitis and hyperthyroidism, and an increased risk of pneumonitis (Supplementary Table 3). This was confirmed by the literature for overall ICI-related irAEs and in particular skin toxicities (Eun et al., 2019; Nakamura et al., 2019; Pavan et al., 2019; Lee et al., 2021). However, other publications did not find that baseline NLR was significantly associated with irAEs (Khoja et al., 2016; Matsukane et al., 2021). Combining NLR and platelet in SII did not add any prognostic value to NLR, and NLR remained the dominant risk factor (Supplementary Table 3).

Several other serum chemistry parameters initially suspected of having a potential correlation with risk of irAEs have not been confirmed. In particular, Valpione et al. (2018) and Yamaguchi et al. (2018) found that LDH and C-reactive protein (CRP) at baseline were not associated with irAEs in patients with melanoma and NSCLC receiving ICIs. Likewise, our results did not show consistent relationships between baseline LDH or CRP across different irAEs.

### Tumor mutational burden

Tumor mutational burden (TMB) has emerged as a potential novel biomarker for response to ICI therapy; however, there have been conflicting findings on the association of TMB with irAEs. While a high number of tumor neoantigens is associated with increased tumor immunogenicity, this has not consistently translated into an increased risk for the subsequent loss of tolerance to healthy tissues. Although there is limited evidence demonstrating high TMB association with increased risk of irAE in the post-approval surveillance setting (Bomze, 2019), several reports from clinical trial experience demonstrate no correlation of TMB with the risk of irAE (Wells et al., 2017; Osipov et al., 2020). Similarly, we did not find that tumor mutational burden (TMB) was significantly associated with irAEs (Figure 2 and Supplementary Table 1). Given almost 67% of patients in our analysis had lung cancer, and clinically these patients are at higher risk of pulmonary complications, we also evaluated any relation between baseline TMB levels and the occurrence of irAE pneumonitis in patients with lung cancer. The results did not demonstrate any significant association.

### Study limitations

In addition to the retrospective nature of our meta-analysis, there are other limitations that should be considered while interpreting the findings. In particular, the impact of concomitant medications, such as corticosteroids and other immunosuppressive therapies, on toxicities from ICI therapy were not analyzed. The broad definitions used to capture the irAE medical concepts and the blinded nature of studies when the safety events were reported could have likely confounded the interpretation of data for risk factors between atezolizumab and comparator arms. Furthermore, we only assessed baseline values; an interesting extension of the analyses could be to evaluate the values prior to and after any ICI infusion such as the monitoring of laboratory parameters after treatment start and prior to the occurrence of an irAE.

### Conclusion

This comprehensive meta-analysis based on data from 10,344 patients enrolled in 15 Roche sponsored clinical trials with atezolizumab across 5 solid tumor indications is one of the largest analyses to date that provides insights into patient-level drivers of irAEs, an area of ongoing investigation. Overall, findings from our analysis corroborates with prior reviews and suggests that the reported rate of irAEs with PD-(L)1 inhibitors are nominally lower than CTLA-4 inhibitors. In our analysis, candidate risk factors were associated with the onset of irAE but the effect size was relatively modest. These results do not necessarily imply that changes to clinical practice with regard to safety monitoring of specific patient populations are needed. In our meta-analysis, ethnic origin was the only relatively strong and consistent risk factor identified across multiple irAEs, with Asians being at higher risk of rash, hepatitis and pneumonitis. Country specific reporting differences may be a significant confounder and limit the interpretation of these findings in Asian patients. However, in line with the principles of inclusive research, there is need for additional data to address outstanding questions to characterize the impact of ethnicity and genetic ancestry on ICI associated immune toxicities.

## Data Availability

Qualified researchers may request access to individual patient level clinical data for each separate study through a data request platform. At the time of writing this request platform is Vivli https://vivli.org/ourmember/roche/. The datasets are only available as individual datasets per study and not integrated across studies.
For up to date details on Roche's Global Policy on the Sharing of Clinical Information and how to request access to related clinical study documents, see here: https://go.roche.com/data_sharing
Anonymised records for individual patients across more than one data source external to Roche can not, and should not, be linked due to a potential increase in risk of patient re-identification.

## Supplementary Table Legends

**Suppl. Tab. 1:** Confounder-adjusted results from the IPD meta-analysis for all tested baseline risk factors (rows). Columns report the results for the association with each irAE type of interest in terms of hazard ratios, 95%-confidence intervals (CI, lower and upper bound), (unadjusted) p-values and FDR-adjusted p-values (p). A separate table sheet is provided for each sub-analysis performed: “Overall” (all indications, all treatment arms and any irAE grade), “Lung c.” (only lung cancer studies, all treatment arms and any irAE grade), “UC” (only UC studies, all treatment arms and any irAE grade), “RCC” (only RCC studies, all treatment arms and any irAE grade), “Atezo arms” (all indications, only atezolizumab arms and any irAE grade), “SOC arms” (all indications, only standard of care arms and any irAE grade), “AE grade 2-5” (all indications and all arms, only grade 2+ irAEs), “AE grade 3-5” (all indications and all arms, only grade 3+ irAEs), “AE grade 3-5 Atezo arms” (all indications, only atezolizumab arms and grade 3+ irAEs). Abbreviations of risk factors: ALP=Alkaline Phosphatase, ALT=Alanine Aminotransferase, aPTT=activated Partial Thromboplastin Time, AST=Aspartate Aminotransferase, BUN=Blood Urea Nitrogen, CRP=C-Reactive Protein, ECOG=Eastern Cooperative Oncology Group performance status, LDH=Lactate Dehydrogenase, NLR=Neutrophil-Lymphocyte Ratio, PD-L1 IC/TC=PD-L1 immunohistochemistry staining intensity scores for immune cells (IC) and tumor cells (TC) (IC neg including values IC0 and neg, IC pos including IC1-3 and pos), SLD=Sum of the Longest Diameters, T3 free=free Triiodothyronine, TMB=Tumor Mutational Burden, TSH=Thyroid Stimulating Hormone.

**Suppl. Tab. 2:** Results of the IPD meta-analysis for the demographic factors used as confounders in the model for each risk factor. This IPD meta-analysis model included the following fixed effects: sex (female vs male), BMI (kg/m2), age (years) and treatment (Atezolizumab-monotherapy, Atezolizumab-combination, standard of care) and study as a random effect. The analysis was performed across all indications, treatment arms and for any irAE grade. Results are reported in terms of hazard ratios, 95%-confidence intervals (CI, lower and upper bound), and (unadjusted) p-values.

**Suppl. Tab. 3:** Confounder-adjusted results from the IPD meta-analysis to assess the association between Neutrophil-Lymphocyte Ratio (NLR) and Platelet either as separate risk factors or in combination (either simultaneous inclusion of both risk factors in the model, or combined in the new variable systemic immune-inflammation index [SII=NLRxPlatelet]) with the onset of irAEs (any grade) of interest. Results are reported in terms of hazard ratios (HR), 95%-confidence intervals (CI, lower and upper bound), and unadjusted/nominal p-values. Additionally, the number of patients (N) and irAEs (Events) is shown, based on the patients without any missing values in the variables included for each model.

## List of Abbreviations

AJ: Aalen-Johansen (estimator)
ALP: alkaline phosphatase
ALT: alanine aminotransferase
AST: aspartate aminotransferase
BMI: body mass index
CI: confidence intervals
CIF: cumulative incidence function
CRP: C-reactive protein
CTCAE: Common Terminology Criteria for Adverse Events
FDR: false discovery rate
HR: hazard ratio
ICI: immune checkpoint inhibitors
IPD: individual-patient data (meta-analysis)
irAE: immune-related adverse events
LDH: lactate dehydrogenase
LFT: liver function test
NLR: neutrophil-lymphocyte ratio
(N)SCLC: (non-)small cell lung cancer
RCC: (advanced) renal cell carcinoma
SD: standard deviation
SII: systemic immune-inflammation index
SL: study-level (meta-analysis)
SLD: sum of the longest diameters
SOC: standard of care
TMB: tumor mutational burden
TNBC: triple-negative breast cancer
TSH: thyroid stimulating hormone
UC: urothelial bladder cancer
ULN: upper limit of normal range

