## Supplementary material for "Baseline risk factors associated with immune related adverse events and atezolizumab": Supp Fig 1

**Suppl Fig 1:** Cumulative incidence function stratified by study (panels) and treatment arm (color) to estimate the irAE probability (any grade) over time while considering death as a competing event. One separate plot for each irAE type: A) Rash, B) Hepatitis, C) Hypothyroidism, D) Hyperthyroidism, E) Pneumonitis.

A) Rash

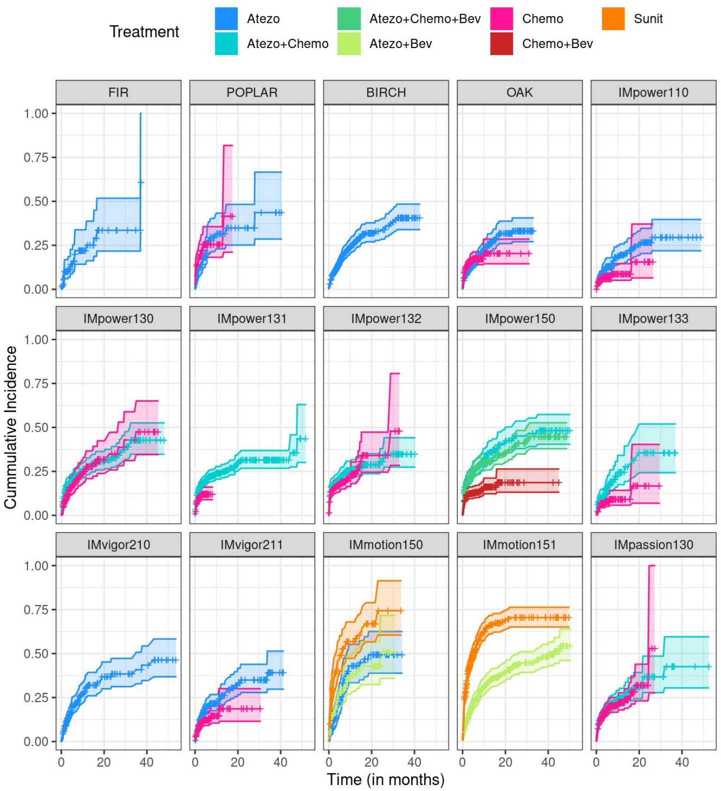


B) Hepatitis

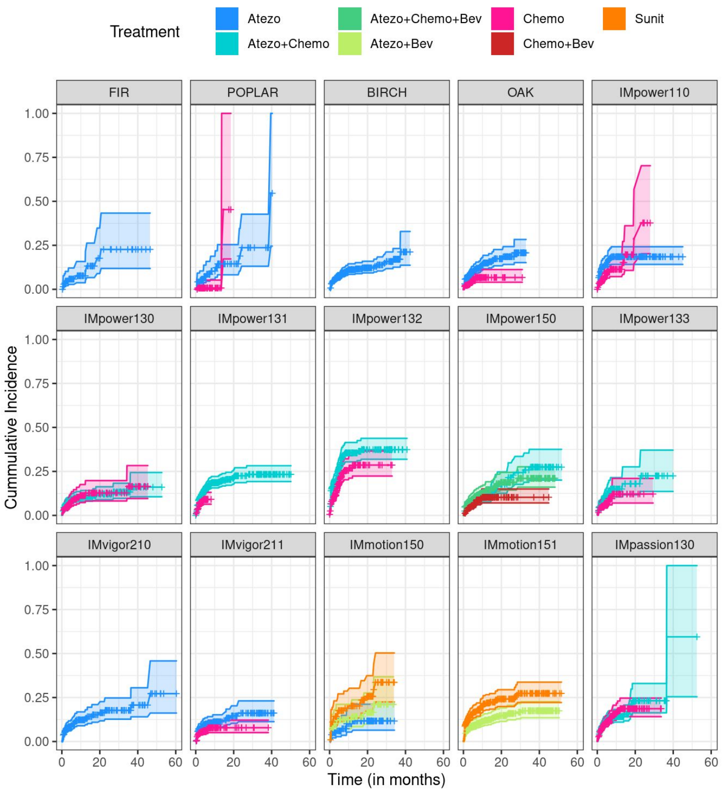

**Suppl Fig 1:** Cumulative incidence function stratified by study (panels) and treatment arm (color) to estimate the irAE probability (any grade) over time while considering death as a competing event. One separate plot for each irAE type: A) Rash, B) Hepatitis, C) Hypothyroidism, D) Hyperthyroidism, E) Pneumonitis.

C) Hypothyroidism

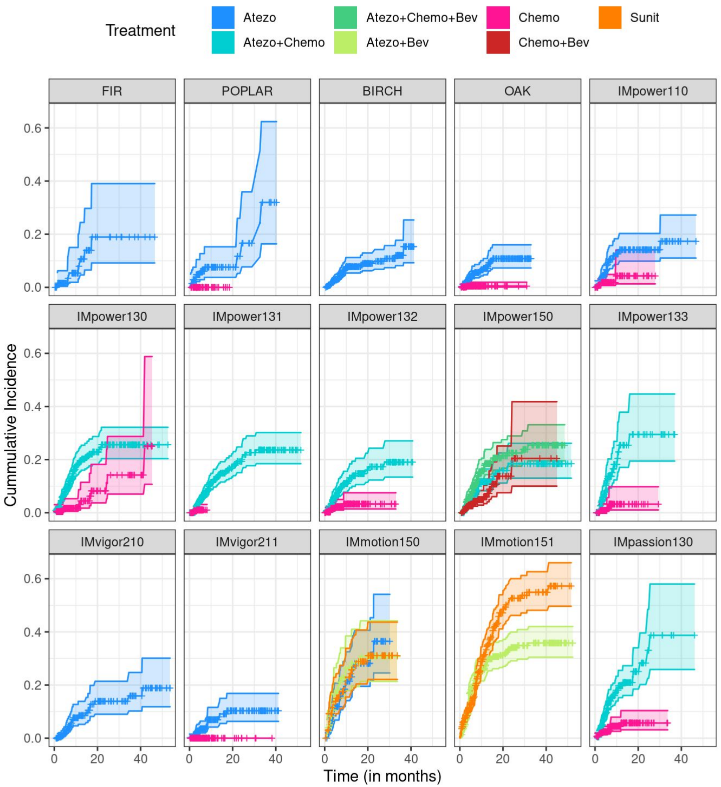


D) Hyperthyroidism

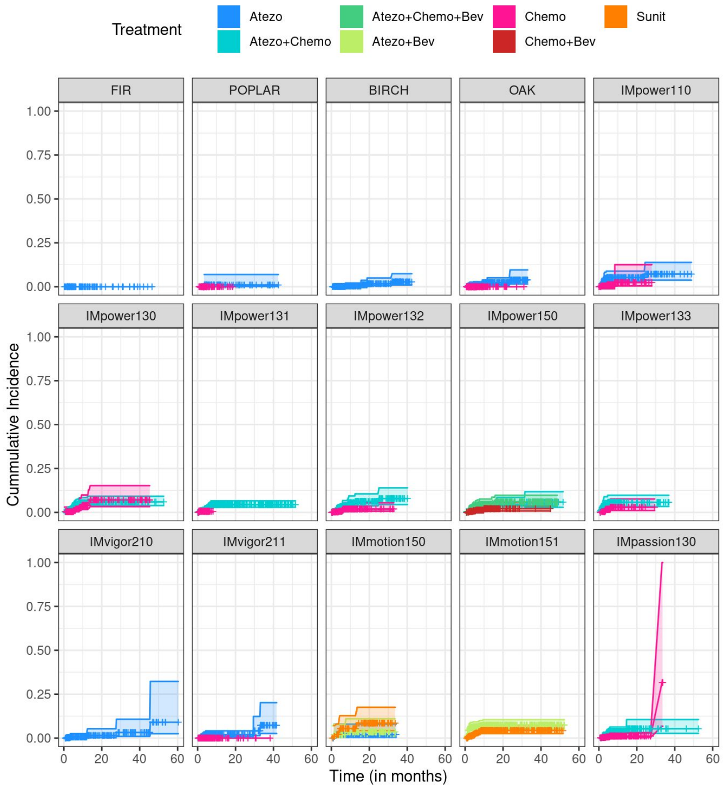

**Suppl Fig 1:** Cumulative incidence function stratified by study (panels) and treatment arm (color) to estimate the irAE probability (any grade) over time while considering death as a competing event. One separate plot for each irAE type: A) Rash, B) Hepatitis, C) Hypothyroidism, D) Hyperthyroidism, E) Pneumonitis.

E) Pneumonitis

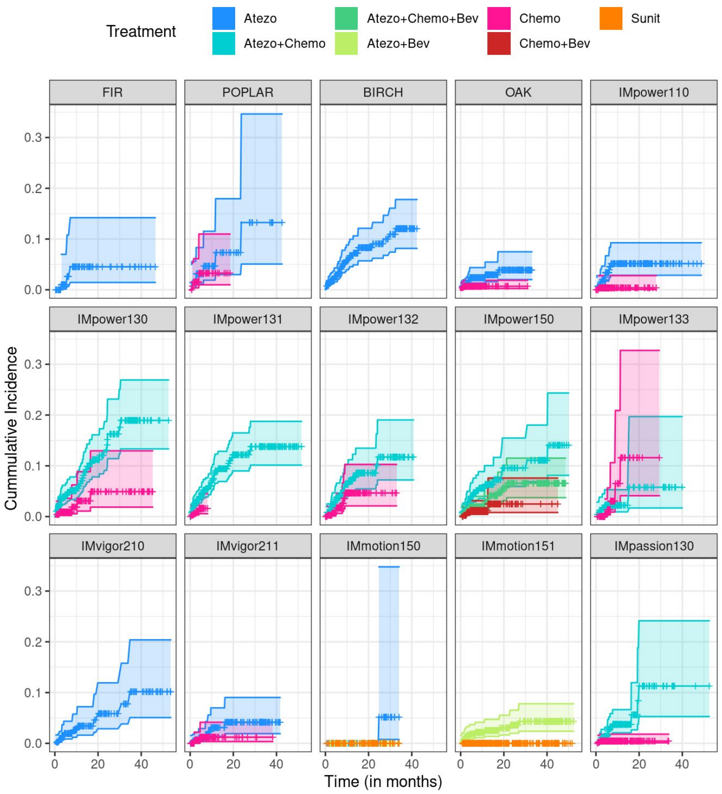
