## Supplementary material for "Baseline risk factors associated with immune related adverse events and atezolizumab": Supp Fig 2

**Suppl Fig 2:** IPD meta-analysis results of the confounding factors age (years), sex (female vs male) and BMI (kg/m2) that were included in every risk factor model. Only confounders exhibiting a consistent and strong association with the selected irAEs of interest are shown. The baseline risk factors listed at the left side of each heatmap correspond to the IPD meta-analysis models where the confounder of interest was significant (FDR p-value < 0.05).

The color of the cells in the heatmap represents the direction of the confounder effect (red: HR>1, i.e. increased irAE risk; blue: HR<1, i.e. decreased irAE risk) and the strength of association (the darker the color the stronger the effect). The symbol within each cell indicates the significance level (FDR p-value <0.1: *, FDR p-value <0.05: **).

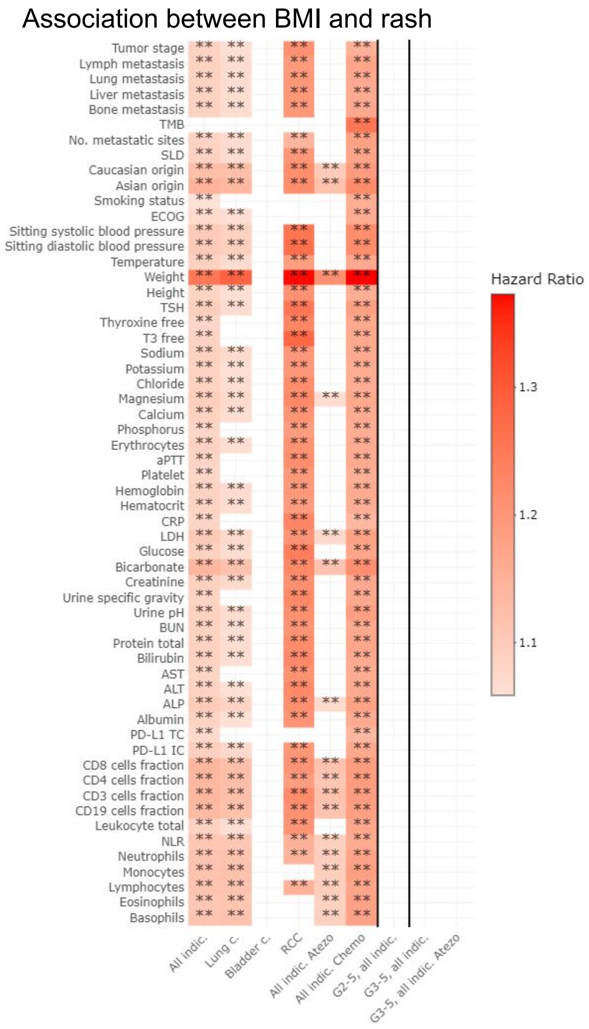

Association between age and hepatitis

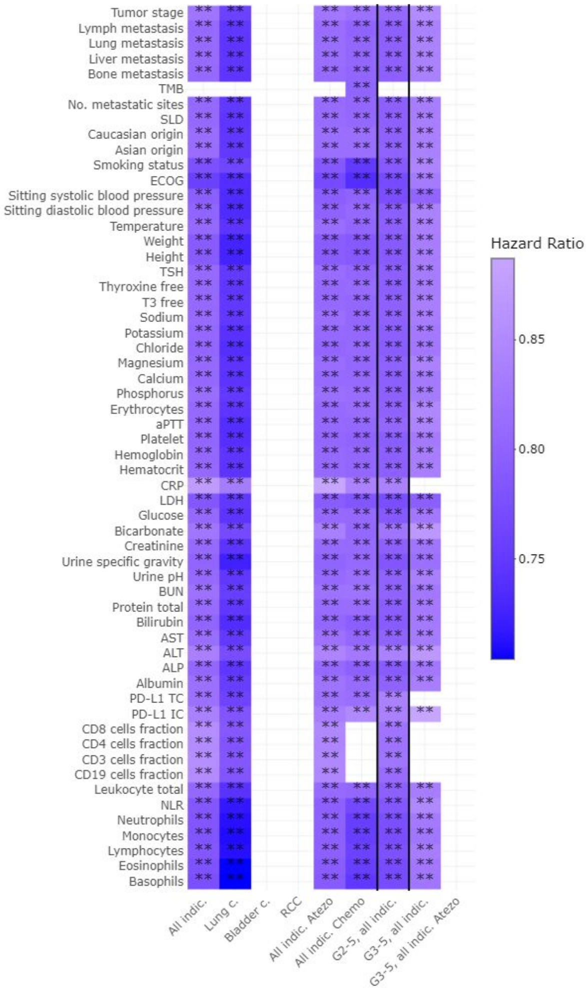


Association between sex and pneumonitis

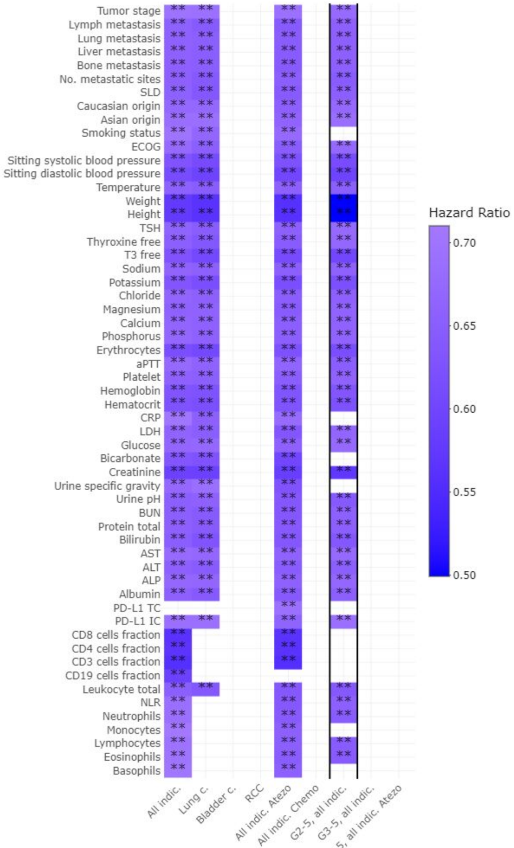


Association between sex and hypothyroidism

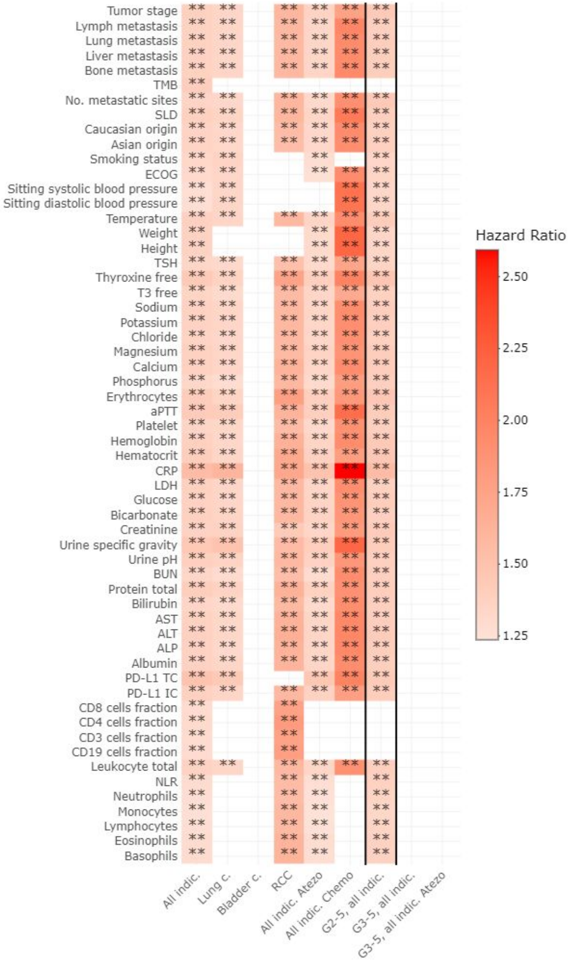
