## Supplementary material for "Baseline risk factors associated with immune related adverse events and atezolizumab": Supp Fig 3

**Suppl Fig 3:** Comparison of the risk factors results (hazard ratios [HR] and confidence intervals [CI]) from the confounder-adjusted individual patient data meta-analysis between the atezolizumab (Atezo) and standard of care (SOC) arms. The color refers to the significance level (FDR adjusted p-values). One separate plot for each irAE type: A) Rash, B) Hepatitis, C) Pneumonitis, D) Hypothyroidism, E) Hyperthyroidism.

A) Rash

B) Hepatitis

C) Pneumonitis

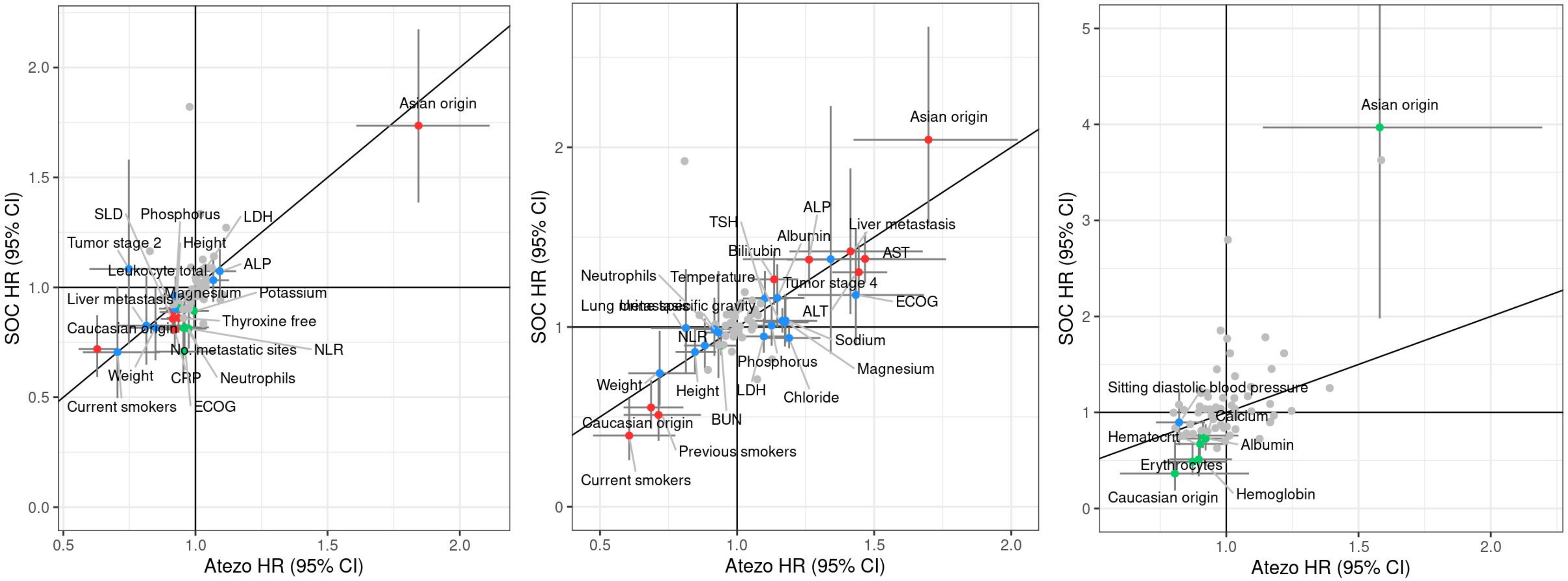

**Suppl Fig 3:** Comparison of the risk factors results (hazard ratios [HR] and confidence intervals [CI]) from the confounder-adjusted individual patient data meta-analysis between the atezolizumab (Atezo) and standard of care (SOC) arms. The color refers to the significance level (FDR adjusted p-values). One separate plot for each irAE type: A) Rash, B) Hepatitis, C) Pneumonitis, D) Hypothyroidism, E) Hyperthyroidism.

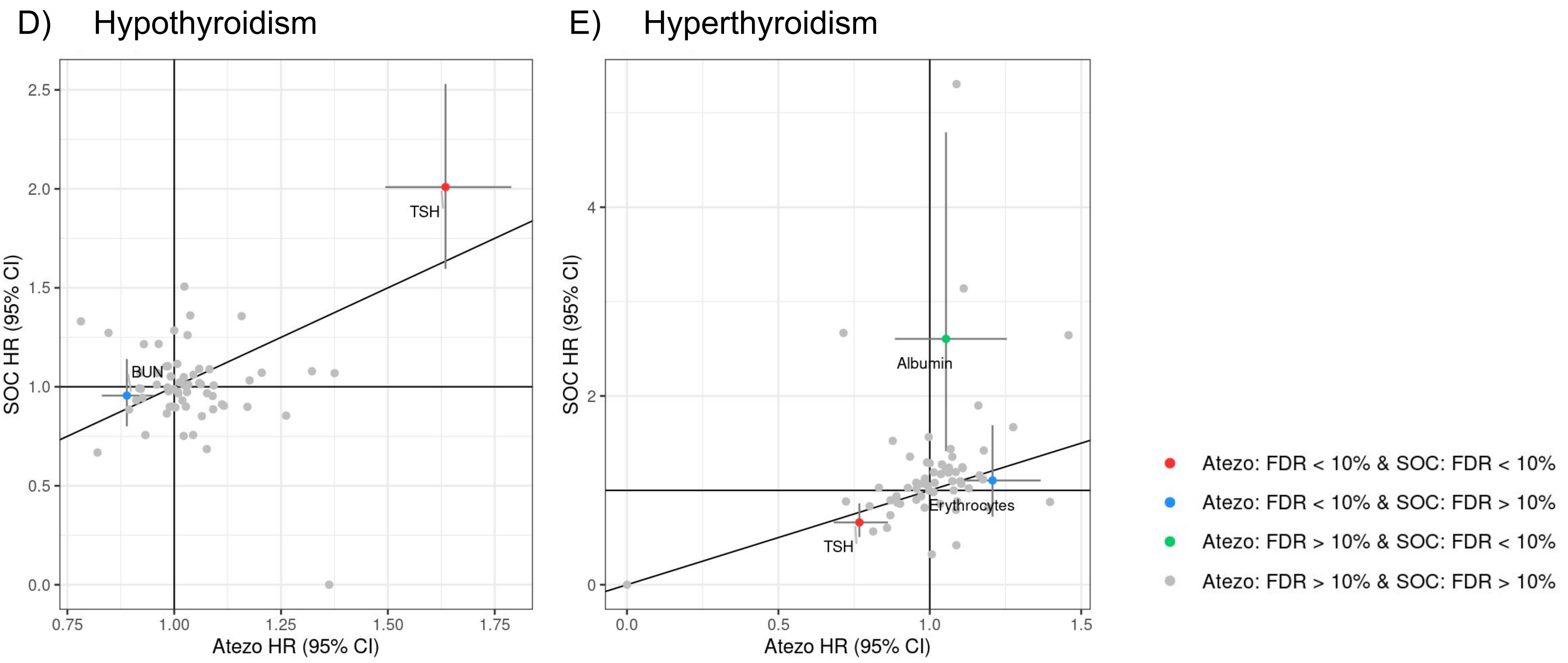
